# Smoking, race, ancestry and prospective abstinence

**DOI:** 10.1101/2021.12.24.21267950

**Authors:** Andrew W Bergen, Carolyn M Ervin, Christopher S McMahan, James W Baurley, Harold S Javitz, Sharon Hall

## Abstract

**Background:** Factors influencing cessation include biopsychosocial characteristics, treatments and responses to treatment. The first cessation trial designed to assess cessation disparities between African American and White cigarette smokers demonstrated that socioeconomic, treatment, psychosocial and smoking characteristics explained cessation disparities. Ongoing translational efforts in precision cessation treatment grounded in genetically informed biomarkers have identified cessation differences by genotype, metabolism, ancestry and treatment.

**Methods:** In planned analyses, we evaluated six smoking-related measures, demographic and socioeconomic covariates, and prospective abstinence (7-day point prevalence at 12 weeks with bupropion, nicotine replacement and counseling treatments). We assessed concurrent and predictive validity in two covariate models differing by inclusion of Office of Management and Budget (OMB) race/ethnicity or genomic ancestry.

**Results:** We studied Pharmacogenetic Study participants (N=456, mean age 49.5 years, 41.5% female, 7.4% African American, 9.4% Multiracial, 6.5% Other, and 6.7% Hispanic). Cigarettes per day (OR=0.95, *P*<.001), Fagerström score (OR=0.89, *P*<=.014), Time-To-First-Cigarette (OR=0.75, *P*<=.005) and predicted urinary nicotine metabolite ratio (OR=0.57, *P*<=.039) were associated with abstinence. OMB African American race (ORs from 0.31 and 0.35, *p-values*<=.007) and African genomic ancestry (ORs from 0.21 and 0.26, *p-values*<=.004) were associated in all abstinence models.

**Conclusions:** Four smoking-related measures exhibited association with abstinence, including predicted nicotine metabolism based on a novel genomic model. African genomic ancestry was independently associated with reduced abstinence. Treatment research that includes social, psychological, treatment and biological factors is needed to reduce cessation disparities.

**Implications:**

- This is the first application of a statistical learning model of the urinary nicotine metabolite ratio to cessation. Results are concordant with biochemical and genetic models of the plasma nicotine metabolite ratio in multiethnic samples.
- The urinary ratio exhibits the largest standardized effect size of four smoking-related measures associated with cessation (time-to-first cigarette, total Fagerström score and cigarettes per day were the others).
- The social construct of African American race and genomic African ancestry are significant covariates in all cessation models.
- Results point to social and biological mechanisms requiring investigation in larger samples to understand and reduce cessation health disparities.

## INTRODUCTION

Tobacco-attributable disease remains the largest modifiable source of mortality in the United States^1^. Numerous tobacco-related health disparities (TRHDs) exist in the United States due to inequities in tobacco related exposures, in social resources supporting healthy behaviors, and in access to health care^2^. Demographic, socioeconomic and health condition TRHDs include greater tobacco-related cancer incidence and mortality in Non-Hispanic Black *vs* White smokers^1^, increased smoking among the less well educated^3^, and excess smoking and related mortality in those with mental illness^4^. Among efforts to reduce these complex TRHDs, better understanding of the factors and mechanisms underlying cessation disparities will support development of more effective interventions.

Nollen *et al* provided evidence on cessation disparities and explanatory factors between African American and White smokers from planned analyses of the first prospective trial designed to test such differences: “Understanding Disparities in Quitting in African American and White Smokers” (NCT01836276^5,6^). In *N*=448 Midwestern smokers of modest income stratified by race, age, and gender, Black smokers were significantly less likely than White smokers to be abstinent (7-day point prevalence at the end of a varenicline and counseling intervention, with Intent-To-Treat abstinence OR=0.51 (95% CI 0.32-0.83) *P<*.007, and in completers only, a OR=0.42 (0.26-0.69) *P<*.001)^6^. In preplanned analyses of age, sex, race and factors associated with race and abstinence, they identified home ownership, income, neighborhood problems, study visits completed, and plasma cotinine in a highly predictive best fit joint model (83% concordance with abstinence). They concluded that the best fit model was supported by pathway analyses linking employment and education to cessation through negative affect, across races^7^.

Nollen *et al* identified cessation disparities and explanatory factors in analyses of US smokers in the randomized placebo-controlled trial called “Evaluating Adverse Events in a Global Smoking Cessation Study” (EAGLES, NCT01456936)^8,9^. EAGLES was designed to test the relative neuropsychiatric risks and efficacy of varenicline and bupropion to those of nicotine replacement therapy (NRT) and placebo; a total of *N*=8144 smokers (>=10 cigarettes per day, CPD) with and without psychiatric disorders were enrolled^9^. The primary outcome was continuous abstinence from weeks 9 - 24 (abstinence). Initial analyses demonstrated no significant increase in neuropsychiatric adverse events due to varenicline or bupropion compared to NRT or placebo, or across diagnostic cohorts; greatest effectiveness was observed with varenicline, and with bupropion and NRT *versus* placebo. In analyses of data from *N*=4109 US EAGLES participants (25.9% Black and 74.1% White)^8^, Nollen *et al* examined treatment group, psychiatric cohort, and race stratified groups for association with abstinence, then examined 41 participant and trial characteristics for association with race and with abstinence, and constructed a best joint model of abstinence. They observed reduced abstinence over all treatments and diagnostic cohorts in Black participants when compared to White participants (OR=0.53 (0.41-0.69) *P<*.001). Similar disparities were observed for varenicline and bupropion arms across cohorts, within the psychiatric cohort, but not within the non-psychiatric cohort. From 19 participant and trial characteristics associated with both race and abstinence, the final abstinence model retained six baseline variables (age, race, anxiety, Fagerström Test for Nicotine Dependence (FTND), CPD, and lifetime quit attempts) and three trial variables (study drug discontinuation, depression score change and emergent adverse events). These analyses have provided evidence that the social construct of race, and demographic, psychometric, smoking, socioeconomic and treatment measures are associated with cessation disparities^8,9^.

Nicotine metabolite biomarkers include the nicotine metabolite ratio (NMR, *trans*-3’-hydroxcotinine/cotinine)^10^, which measures metabolism, and total nicotine equivalents (TNE, the molar sum of nicotine and its principal metabolites including cotinine, COT, and *trans*-3’-hydroxycotinine, 3HC), which measures intake^11^. The plasma NMR and the urinary NMR are associated with CPD^12,13^, and TNE is associated with the FTND and its CPD and Time-To-First-Cigarette items^14^. The NMR, and another genetically-informed biomarker of variants at the alpha 5 nicotinic acetylcholine receptor gene (*CHRNA5*) associated with CPD, cessation and lung cancer^15–17^, are being translated into precision cessation therapy^18^. A prospective stratified randomized trial of the NMR and of varenicline, NRT and placebo, with primary abstinence outcome 7-day point prevalence abstinence at end of treatment, identified a NMR-by-treatment interaction (NCT01314001)^19^. Normal nicotine metabolizers achieved greater abstinence with varenicline, while slow metabolizers achieved equivalent abstinence with either varenicline or NRT but reported greater adverse events with varenicline, suggesting a treatment strategy for translational research. A prospective stratified randomized trial of the *CHRNA5* biomarker and of varenicline, combined NRT (patch and lozenge) and placebo with primary abstinence outcome 7-day point prevalence abstinence at end of treatment identified a genotype-by-treatment interaction within non-European American (88% African American) participants (NCT02351167)^20^. Those participants with higher risk genotypes (rs16969968 GA/AA), achieved greater abstinence with varenicline *vs* placebo, while participants with the low risk genotype (rs16969968 GG) achieved greater abstinence with cNRT *vs* placebo.

African American and European American cigarette smokers differ in nicotine metabolite and smoking intensity measures; African American smokers exhibit lower levels of CPD, exhaled carbon monoxide (CO), the NMR (plasma and urinary), TNE and plasma COT+3HC, but higher plasma COT and higher nicotine exposures per cigarette^21–23^. Differences in nicotine metabolites were hypothesized to contribute to TRHD^21^. Robust associations of nicotine and toxicants in Black and White smokers^24^, modeling smoking intensity *vs* total nicotine equivalents and risk for lung cancer in multiethnic populations^25^, and increased toxicant exposures in African American smokers^26^, support the hypothesis that increased tobacco exposures per cigarette contribute to TRHD.

In this study, we performed planned analyses to evaluate the utility of four self-reported smoking measures (cigarettes per day, CPD; the FTND CPD item, FCPD; time-to-first-cigarette, TTFC; and total FTND score, FTND) and two biomarkers (exhaled carbon monoxide, CO, and predicted urinary nicotine metabolism, uNMR), to predict prospective abstinence in participants recruited from two randomized clinical trials^27,28^. We assessed association of these six smoking-related measures with demographic, socioeconomic and ancestry covariates, and then with prospective abstinence. We estimated and compared the influence of two covariate models, one containing the social constructs of race and ethnicity and the other, genomic ancestry.

## METHODS

### Ethical Approval

Written informed consent was obtained from all participants. Research described herein received approvals from the Institutional Review Boards of BioRealm and Oregon Research Institute.

### Participants (N=456)

We studied existing clinical and novel genomic data from participants of two randomized smoking cessation trials (RCT NCT00087880^28^ and RCT NCT00086385^27^, who provided whole blood and consent for a study called Pharmacogenetics of Bupropion and Nicotine Smoking Cessation Trial (Pharmacogenetic Study)^29^ (**Table 1)**. See **Supplementary Materials** for additional details on the RCTs and Pharmacogenetic Study.

**Table 1:** Pharmacogenetic Study Participant Characteristics, N=456

| Characteristic | Definition | Type | N (%) or M (SD) |
| --- | --- | --- | --- |
| RCT Source | NCT00087880 | Clinical | 220 (48.2%) |
| Taper | Taper NRT | Clinical | 112 (24.6%) |
| Session | 7 Sessions | Clinical | 96 (21.1%) |
| Age | <50 years | Demographic | 187 (41.0%) |
| Gender | Female | Demographic | 189 (41.5%) |
| Education | <College graduate | Social | 224 (49.3%) |
| African American | Self-identified | Social | 33 (7.4%) |
| Multiple | Self-identified | Social | 42 (9.4%) |
| Other | Aggregated* | Social | 29 (6.5%) |
| White | Self-identified | Social | 343 (76.7%) |
| Hispanic | OMB Ethnicity | Social | 30 (6.7%) |
| BMI <25 kg/m<sup>2</sup> | Adiposity | Clinical | 203 (45.7%) |
| African Ancestry | Continuous | Genomic | 0.098 (0.265) |
| Asian Ancestry | Continuous | Genomic | 0.076 (0.224) |
| CO (ppm) | Carbon monoxide | Smoking | 20.9 (10.8) |
| CPD | Cigarettes per Day | Smoking | 19.4 (7.7) |
| FTND (0-10) | Fagerström Test** | Smoking | 4.76 (2.1) |
| TTFC (0-3) | Time-To-First-Cigarette | Smoking | 1.84 (0.98) |
| FCPD (0-3) | Fagerström CPD | Smoking | 1.44 (0.68) |
| uNMR | Predicted uNMR | Genomic | 1.22 (0.43) |
| Abstinent | Completor only | Clinical | 265/429 (61.8%) |
| Abstinent (ITT) | By ITT method | Clinical | 265/456 (58.1%) |

\*Modified Other (Asian, Native American, Other, Pacific Islander). \*\*for Nicotine Dependence.

### Pharmacogenetic Study Recruitment

This study was initiated when the RCTs were in progress. Thus, cessation trial participants were approached at their first scheduled RCT assessment after the initiation of the present study, and were asked if they would be willing to participate in a study of the genetics of response to smoking cessation treatment with nicotine and bupropion, dependence on tobacco smoking, and nicotine metabolism and clearance. The Pharmacogenetic Study was described, participants read the consent form, were given an opportunity to ask questions, and interested participants signed the consent form. Participants were paid $50 for providing a blood sample.

### Genomic Prediction of Ancestry and uNMR

Pharmacogenetic Study participant DNA was prepared from whole blood (Gentra Puregene, Qiagen, Germantown, MD), and genotyped at Infinite Biologics (Piscataway, NJ). Genotype data was subjected to quality control and genotype analysis.

Ancestry proportions were estimated by extracting genotypes from N=5,516 ancestry informative markers from the Pharmacogenetic Study data. After combining these genotypes with genotypes from 1000 Genomes Project Phase 3 version 5a, fastSTRUCTURE was used with default settings and *k*=3 populations^30^. Population assignments from the 1000 Genome Project and self-reported race were used to label estimated African, Asian, and European ancestry proportions.

We predicted the natural log transformed urinary ratio of total 3HC/COT (uNMR_*T /F*_, hereafter uNMR) in Pharmacogenetic Study participants using participant clinical and genotype data based on an ensemble model trainedon Multiethnic Cohort (MEC) data^31^. In brief, the ensemble model was created using MEC participant age, gender, BMI, genomic ancestry and variants nominated from four genome-wide association scans of the NMR to train seven statistical models using measured MEC uNMR data^32^. The average of the seven models is the ensemble estimate.

For additional details, see **Supplementary Materials**.

### Clinical Treatment Variables

Variables indicating source RCT^27,28^, instructions to taper from NRT from weeks 8 to 12^27^, and assignment of additional counseling sessions in weeks 10 and 12^27^, were created to address variations in treatments across trials, and among participants in the last month before the 12 week assessment.

### Imputation of Missing Data

Demographic and social categorical covariates were stratified (reference) for analysis: age (*<*50 years); gender (female); education (*<*college graduate); modified OMB race with African American, Multiple, Other (grouping four infrequent categories: Asian, Native American, Pacific Islander, and Other), and (White); OMB ethnicity (Non-Hispanic); and BMI (*<*25 kg/m^2^). Missing data were estimated using the missMDA R package with the estim ncpFAMD and imputeFAMD functions^33^. Abstinence was assigned using the intent-to-treat (ITT) approach. All multivariate analyses were performed using the N=456 post-imputation dataset.

### Statistical Analyses

We stratified participants by prospective abstinence and by covariates for descriptive analyses. We performed concurrent validity analysis of the predicted uNMR with linear regression with smoking measures as the outcomes. We used two covariate models in analysis. Covariate Model 1 included age, gender, education, OMB race, OMB ethnicity, and BMI, and Covariate Model 2 included age, gender, education, ancestry proportions and BMI. Predictive validity of smoking measures and uNMR with abstinence was assessed with logistic regression and the same covariate models. Descriptive analyses were performed in JMP (v. 15.0.0, SAS Institute Inc., Cary, NC, USA) and multivariate analyses in the R statistical language. The α level was .05 and tests of significance were two-sided.

## RESULTS

Pharmacogenetic Study participant recruitment did not differ by RCT, and Pharmacogenetic Study participant demographic, social and smoking characteristics did not differ from RCT participant characteristics. See **Supplementary Table 1** for RCT and Pharmacogenetic Study recruitment numbers, **Supplementary Table 2** for comparison of RCT and Pharmacogenetic Study characteristics, and **Supplementary Materials** for details on RCT eligibility and interventions.

### Univariate Analysis

Data missingness was low: demographic and social covariates, genomic variables, and smoking measure variable missingness rates were 0.0% - 2.0%, 4.0%, and 0.0% - 2.4%. Clinical variables were not associated with abstinence (*p-values>=*.50), and were not included in further analyses. Education (*P*=.096), African American OMB race (*P*=.012), African ancestry (*P*=.002), CPD (*P*=.007), TTFC (*P*=.003), FTND score (*P*=.010) and the uNMR (*P*=.052) were significantly associated with abstinence (**Supplementary Table 3**). The directionality of education and NMR associations with abstinence are known, thus these associations were considered significant. All covariates except Hispanic ethnicity were significantly associated with CPD, FCPD and/or uNMR while gender and education were associated with the FTND (**Table 2**). There were no covariate associations with the CO or TTFC smoking measures. Participants who were younger, female, college-educated, minority OMB race or ethnicity, normal weight or with African ancestry or Asian ancestry were more likely to exhibit reduced smoking measure levels, while covariate relations with uNMR were mixed. See **Supplementary Table 4** for stratified covariates.

**Table 2:** Sociodemographics, Smoking Measures and uNMR

|  |  | CO | CPD | FTND | TTFC | FCPD | uNMR |
| --- | --- | --- | --- | --- | --- | --- | --- |
| Age | *t* | 1.05 | 3.28 | 0.313 | 1.54 | 3.29 | 1.30 |
|  | *P* | .293 | .001 | .754 | .125 | .001 | .195 |
| Gender | *t* | -1.28 | -4.12 | -2.04 | -1.11 | -2.74 | 1.53 |
|  | *P* | .201 | <.001 | .042 | .269 | .006 | .127 |
| Education | *t* | -1.23 | -1.93 | -2.15 | -1.72 | -0.255 | 1.15 |
|  | *P* | .221 | .055 | .032 | .087 | .799 | .249 |
| African American | *t* | -0.11 | -2.45 | -0.87 | -0.59 | -2.18 | -2.58 |
|  | *P* | .914 | .015 | .39 | .557 | .030 | .010 |
| Multiple | *t* | -1.04 | -1.63 | -1.27 | -1.08 | -2.20 | -4.71 |
|  | *P* | .299 | .103 | .205 | .281 | .029 | <.001 |
| Other | *t* | -1.16 | -2.18 | -0.92 | -0.10 | -2.80 | -5.79 |
|  | *P* | .247 | .030 | .359 | .923 | 0.005 | <.001 |
| Hispanic | *t* | 1.06 | 0.142 | 1.28 | 0.724 | 0.726 | -0.317 |
|  | *P* | .290 | .887 | .202 | .469 | .468 | .751 |
| BMI | *t* | 0.03 | 1.74 | 1.29 | 0.763 | 1.87 | -2.62 |
|  | *P* | .979 | .083 | .199 | .446 | .063 | .009 |
| African ancestry | *t* | -0.58 | -2.27 | -0.296 | 0.675 | -2.06 | -2.73 |
|  | *P* | .561 | .024 | .767 | .500 | .040 | .007 |
| Asian ancestry | *t* | -0.82 | -2.74 | -1.89 | -1.54 | -3.34 | -8.17 |
|  | *P* | .412 | .006 | .060 | .124 | <.001 | <.001 |

Covariates are predictors (reference for categorical variables) and smoking measures are outcomes: Age (<50 years); Gender (Male); Education (<College graduate); OMB Race, African American, Multiple, Other and (White); OMB Ethnicity (Hispanic); BMI (<25) kg/m<sup>2</sup>), African ancestry, Asian ancestry. Smoking Measures: CO (ppm), CPD=Cigarettes per day (continuous), FTND=FTND Total, TTFC=FTND Time-To-First-Cigarette item, FCPD=FTND CPD item. uNMR=Predicted natural log uNMR.

### Concurrent Validity of the uNMR

The uNMR was significantly associated with CO in each covariate model (**Table 3**). Multiple covariates were significantly associated with the smoking measures CPD, FCPD and FTND (**Table 3**). In Covariate Model 1 (**Supplementary Table 5**), age, gender, education, and African American OMB race were associated with CPD, and age, gender, African American OMB race, Other OMB race and BMI were associated with FCPD. In Covariate Model 2 (**Supplementary Table 6**), age, gender, education, African ancestry and Asian ancestry were associated with CPD, and age, gender, African ancestry, Asian ancestry and BMI were associated with the FCPD; also, education and Asian ancestry were associated with FTND.

**Table 3:** uNMR and Smoking Measures

| | $\beta$ | SE | *P* | $r^2$ | adj. $r^2$ | *F P-value* |
| --- | --- | --- | --- | --- | --- | --- |
| Covariate Model 1 |  |  |  |  |  |  |
| CO | 3.80 | 1.29 | .003 | 0.035 | 0.015 | .070 |
| CPD | 1.39 | 0.89 | .120 | 0.099 | 0.081 | <.001 |
| FTCD | 0.18 | 0.25 | .469 | 0.035 | 0.016 | .063 |
| TTFC | 0.10 | 0.12 | .400 | 0.023 | 0.003 | .317 |
| FCPD | 0.07 | 0.08 | .390 | 0.084 | 0.066 | <.001 |
| Covariate Model 2 |  |  |  |  |  |  |
| CO | 3.88 | 1.32 | .004 | 0.032 | 0.017 | .044 |
| CPD | 0.82 | 0.90 | .364 | 0.110 | 0.096 | <.001 |
| FTCD | 0.06 | 0.25 | .830 | 0.036 | 0.021 | .022 |
| TTFC | 0.07 | 0.12 | .585 | 0.023 | 0.008 | .163 |
| FCPD | 0.02 | 0.08 | .815 | 0.096 | 0.082 | <.001 |

The uNMR is the predictor and smoking measures are the outcomes. Covariate Model 1 adjusted for age, gender, education, OMB race and ethnicity, and BMI. Covariate Model 2 adjusted for age, gender, education, ancestry proportions, and BMI.

### Predictive Validity Analyses

Four smoking-related measures exhibited association with abstinence (**Table 4**). Odds ratios (OR (95%CI) *P*) ranged from 0.56 (0.34-0.92) .023 and 0.58 (0.35-0.97) .039 for the uNMR in Covariate Models 1 and 2, to 0.95 (0.93-0.98) *<*.001 for CPD in both Covariate Models. Each individual smoking-related measure had essentially the same standardized effect estimate in both Covariate Models. African American OMB race (Covariate Model 1) and African ancestry (Covariate Model 2) were significantly (*P*<.001) associated with abstinence in all models, including the non-significant FCPD and CO models (**Supplementary Tables 7** and **8**). The reduced odds ratios of abstinence associated with African American OMB race and African ancestry were lower than the odds ratios associated with the CPD and FTND measures (Table 4). The odd ratios of abstinence associated with African ancestry were one-third lower than those associated with African American OMB race, but confidence intervals substantially overlapped. The other covariate associated with abstinence was BMI (*P*=.048) in the CPD model in Covariate Model 2.

**Table 4:** Smoking, OMB Race, Ancestry and Abstinence

| | $\beta$ | SE | *P* | OR | LCI - UCI |
| --- | --- | --- | --- | --- | --- |
| Covariate Model 1 |  |  |  |  |  |
| CO | -0.01 | 0.01 | 0.193 | 0.99 | 0.97 - 1.01 |
| African American | -0.98 | 0.40 | 0.013 | 0.37 | 0.17 - 0.81 |
| CPD | -0.05 | 0.01 | <0.001 | 0.95 | 0.93 - 0.98 |
| African American | -1.16 | 0.40 | 0.004 | 0.31 | 0.14 - 0.69 |
| FTND | -0.12 | 0.05 | 0.012 | 0.89 | 0.81 - 0.97 |
| African American | -1.05 | 0.40 | 0.008 | 0.35 | 0.16 - 0.76 |
| TTFC | -0.31 | 0.10 | 0.003 | 0.73 | 0.60 - 0.90 |
| African American | -1.05 | 0.40 | 0.009 | 0.35 | 0.16 - 0.77 |
| FCPD | -0.29 | 0.15 | 0.057 | 0.75 | 0.56 - 1.01 |
| African American | -1.07 | 0.40 | 0.007 | 0.34 | 0.16 - 0.75 |
| uNMR | -0.58 | 0.25 | 0.023 | 0.56 | 0.34 - 0.92 |
| African American | -1.08 | 0.40 | 0.007 | 0.34 | 0.15 - 0.74 |
| Covariate Model 2 |  |  |  |  |  |
| CO | -0.01 | 0.01 | 0.171 | 0.99 | 0.97 - 1.01 |
| African ancestry | -1.36 | 0.46 | 0.003 | 0.26 | 0.10 - 0.64 |
| CPD | -0.05 | 0.01 | <0.001 | 0.95 | 0.93 - 0.98 |
| African ancestry | -1.57 | 0.47 | <0.001 | 0.21 | 0.08 - 0.53 |
| FTND | -0.12 | 0.05 | 0.014 | 0.89 | 0.81 - 0.98 |
| African ancestry | -1.41 | 0.47 | 0.003 | 0.24 | 0.10 - 0.61 |
| TTFC | -0.29 | 0.10 | 0.005 | 0.75 | 0.61 - 0.92 |
| African ancestry | -1.35 | 0.47 | 0.004 | 0.26 | 0.10 - 0.65 |
| FCPD | -0.28 | 0.15 | 0.063 | 0.75 | 0.56 - 1.02 |
| African ancestry | -1.46 | 0.47 | 0.002 | 0.23 | 0.09 - 0.58 |
| uNMR | -0.54 | 0.26 | 0.039 | 0.58 | 0.35 - 0.97 |
| African ancestry | -1.49 | 0.47 | 0.002 | 0.22 | 0.09 - 0.57 |

Smoking measures are predictors and abstinence is the outcome. Covariate Model 1 adjusted for age, gender, education, OMB race and ethnicity, and BMI. Covariate Model 2 adjusted for age, gender, education, ancestry proportions, and BMI.

## DISCUSSION

This study is the first to relate prospective abstinence and several traditional smoking measures to a predicted measure of nicotine metabolism based on a novel ensemble model we have developed using genomic ancestry. This study is the first to compare the predictive utility of smoking measures in covariate models that differ by inclusion of the social construct of race and genomic measures of ancestry.

### Validation of the predicted uNMR measure

From prior research in both population cohorts and treatment-seeking smokers of multiple ethnicities, with biochemical measures of the NMR and uNMR, and genetic models of the NMR or *CYP2A6* activity, we expected to observe associations between the predicted uNMR and gender, OMB race, BMI, measures of smoking intensity (but not the FTND), and abstinence^13,34–37^. We observed expected associations of the uNMR with OMB race categories and BMI, and novel associations with African ancestry and Asian ancestry proportions in treatment seeking smokers. We observed in multivariate analyses that covariates were significantly associated with measures of smoking intensity (CPD and FCPD) and nicotine dependence (FTND), but not with predicted uNMR. The variables gender, OMB race, BMI and CPD have previously been associated with the NMR in treatment-seeking smokers with similar explained variance^35^ to the associations observed with smoking intensity measures. We did not expect to observe association of the uNMR with CO as a larger sample of treatment-seeking smokers (*N*=1700, 35% African American) did not observe association of the NMR with CO^22^.

The uNMR abstinence association we observed is directionally consistent with previous NMR abstinence research^38^, and has a similar effect size found in a meta-analysis of abstinence at six months in *N*=718 non-Hispanic Black and non-Hispanic White smokers randomized to active NRT comparing smokers with normal NMR to those with slow NMR (Relative Risk of 0.54 and 95%CI 0.37 to 0.78)^39^. Larger samples are necessary to investigate more complex relations (interactions) between the predicted uNMR, covariates, smoking measures and treatment outcomes not tested in our planned analysis.

### Smoking measures, OMB race and ancestry

African American smokers exhibit lower levels of most smoking intensity measures and nicotine biomarkers, except for higher levels of biomarkers of nicotine exposure per cigarette and higher levels of plasma COT; the mechanistic reasons for the first exception are not understood, while the second exception is due to increased NIC intake per cigarette, reduced NIC oxidative metabolism and to reduced NIC and COT glucuronidation^21,40^. The associations we observed in the Pharmacogenetic Study between African American OMB race, decreased smoking intensity measures and predicted nicotine metabolism are consistent with prior studies. Multiple (self-identified OMB multiracial race category) and Other (an aggregated category of self-identified OMB Asian, Native American, Pacific Islander and Other race categories) OMB race categories were associated with reduced smoking intensity and nicotine metabolism measures in the Pharmacogenetic Study, although associations with smoking intensity moderated upon adjustment for nicotine metabolism in multivariate analysis. The reduced smoking measures seen in the aggregated Other category used in this analysis are consistent with reduced CPD and nicotine metabolism in Native Hawaiian and Japanese American smokers^23^.

African ancestry and Asian ancestry were associated with decreased smoking intensity and nicotine metabolism measures in Pharmacogenetic Study participants. Ancestry within self-identified ethnic strata was examined for association with CPD in non-treatment seeking ever smokers from a health maintenance organization (mean(SD) 55(11) years of age and 58% female)^41^. Within group association of the first two geographically interpretable principal components of genetic variation was observed in non-Hispanic White (*N*=12898, 13% current), in East Asian (*N*=921, 15% current), but not in African American (*N*=554, 24% current) ever smokers. However, this analysis differs from our analysis in stratifying by self-identified ethnicity before ancestry analysis and by analyzing ever smokers *vs* treatment-seeking current smokers.

### OMB race and abstinence

Analyses of multiethnic randomized cessation trials have observed an approximately 50% reduction of prospective abstinence among African American smokers in the United States. Therefore, we expected to observe a significant reduction in abstinence in African American smokers in the Pharmacogenetic Study. The range of standardized effect estimates we observed, from ORs (95%CI) of 0.31 (0.14-0.69) in the CPD model to 0.34 (0.15-0.74) in the uNMR model, encompass estimates from studies with a variety of designs: randomized on gender and OMB race^42^, stratified by OMB race^6^, or stratified by psychiatric diagnosis^8^.

### Ancestry and abstinence

One published study identified association of measured African genomic ancestry with prospective abstinence (7-day point prevalence at 4 weeks), using a novel genetic analysis of ancestry and secondary analysis of a cessation trial of naltrexone *vs* placebo where all participants also received NRT and counseling^43^. Bress *et al* observed significantly increased abstinence rates in *N*=136 self-identified White, European ancestry participants randomized to naltrexone but this treatment effect was not observed in *N*=95 self-identified African American, African ancestry participants. Within the African American participants, treatment was associated with increased abstinence in those with *<*80% West African ancestry, while participants with >=80% West African ancestry did not report an effect of treatment. While the European and African ancestry treatment interaction was non-significant, the West African ancestry interaction (*P*<.03) suggested a potential influence of ancestry on response to naltrexone^43^. The findings of Bress *et al* and our findings of African ancestry association with abstinence are not exactly comparable due to differences in trial characteristics and analysis designs. The additional 10% reduction in the odds of abstinence associated with African ancestry compared to African American OMB race in the Pharmacogenetic Study suggests that African ancestry may convey additional reductions in abstinence. Future studies will need to assess potential confounding between ancestry and socioeconomic factors associated with reductions in abstinence in African Americans^6^, and potential moderation, e.g., by gender^44^.

### Translational Implications

Confirmation of the African American TRHD of reduced abstinence using both the social construct of race and genomic ancestry, and the novel association of the predicted uNMR with abstinence, supports future analyses of cessation trials of sufficient sample size to observe and dissect disparities by ancestry^45^, models of nicotine metabolism and consumption^25,46^, socioeconomic factors^6^, and treatments. Such combined biopsychosocial cessation research would support reenvisioning of interventions in the context of the public health crises in the United States^47^.

### Limitations

In this analysis we approached variable analysis in a conservative manner (pre-planned analyses and dichotomous stratification of most covariates), and future research might use alternative approaches. Socioeconomic or related covariates were limited to education or BMI. The most profound limitation was lack of information on use of mentholated cigarettes, subject to targeted marketing to African Americans, and associated with reduced quit ratios in African Americans^48^. For detailed strengths and limitations, please see **Supplementary Materials**.

## Data Availability

The datasets generated during and/or analyzed during the current study are available from the corresponding author on reasonable request.

## Funding

This work was supported by the National Institute on Alcohol Abuse and Alcoholism (R44 AA027675 to JWB and AWB). The sponsor had no role in the analysis of data, writing of the report, or in the decision to submit the paper for publication.

## Declaration of Interests

AWB is an employee of Oregon Research Institute and ORI Community and Evaluation Services, and serves as a Scientific Advisor and Consultant to BioRealm LLC. CME and JWB are members and employees of BioRealm LLC. JWB, CSM and AWB are co-inventors on a related patent application “Biosignature Discovery for Substance Use Disorder Using Statistical Learning”, assigned to BioRealm, LLC. BioRealm LLC offers genotyping and data analysis services. Other authors declare that they have no competing interests.

## Acknowledgements

We thank the participants, Principal Investigators, co-Investigators and staff of the RCTs and the Pharmacogenetic Study. The National Institute on Drug Abuse funded the RCTs (R01 DA02538, K05 DA016752, K23 DA018691, and P50 DA09253), and the Pharmacogenetic Study (U01 DA020830). We thank Denise Nishita, Lisa Jack and Gary Swan (site-PI), Pharmacogenetics of Nicotine Addiction Treatment Consortium at SRI International (U01 DA020830, MPI: NL Benowitz, RF Tyndale and C Lerman), for their essential contributions that enabled this study. We acknowledge the BioRealm LLC team for supporting project workflows and computation, and IBX for DNA sample processing.

## Notes

### Clinical Trial

NCT00087880, NCT00086385

### Author Declarations

IRB of BioRealm waived ethical approval (met the criteria for exemption) for this work. IRB of Oregon Research Institute waived ethical approval (met the criteria for exemption) for this work.

