## Supplementary Materials for "Smoking, race, ancestry and prospective abstinence"

**4 Tables** **15**

**5 References** **24**

7 Covariate Model 1, Smoking Measures, uNMR and Abstinence 22

8 Covariate Model 2, Smoking Measures, uNMR and Abstinence 23

### 1. Chronic Treatment Trials

#### *1.1. Cessation Trial Recruitment*

Participants were recruited at a urban University-associated cessation clinic in the Western US, by diverse local media, provider referral and self-referral. Shared inclusion criteria were ≥10 cigarettes per day (CPD) and intention to quit. Additional criteria were: age ≥18 years, Time-to-First-Cigarette (TTFC) (Heatherton et al. 1991) ≤30 minutes, and regular smoking duration ≥5 years for NCT00087880, and age ≥50 years for NCT00086385. Shared exclusion criteria (see individual RCT exclusion criteria below) included cardiovascular disease, psychiatric disorders, psychiatric medication use, alcohol dependence treatment, non-tobacco substance use disorder, medical and psychiatric histories contraindicated for trial medication, language and physical limitations. At initial contact, the trials were explained in detail, potential participants read the consent forms, given an opportunity to ask questions, and signed the consent forms. Participants were paid $25 per completed assessment.

#### *1.2. Exclusion Criteria*

Exclusion criteria for NCT00087880 included: history of seizure or head injury resulting in unconsciousness; medical condition that might predispose to seizures, e.g., stroke or current/history of anorexia or bulimia; imminently life threatening disease; use of a protease or monoamine oxidase inhibitor within the last two weeks; current use of psychiatric drugs that would interfere with interpretation of study results, including antidepressants; treatment for alcohol dependence in the last year or evidence of severe alcohol abuse; suicidal or homicidal ideation; current major depressive disorder; history of bipolar disorder; recent myocardial infarction, any other medical condition that would contraindicate use of NRT or bupropion; physical impairment so severe that the patient is unable to participate in a program of moderate physical activity; pregnancy or lactation, knowledge that the patient is leaving the study area; and lack of spoken English ability.

Exclusion criteria for NCT00086385 included: history of severe cardiovascular disease; medical condition that might predispose to seizures, e.g., stroke or current/history of anorexia or bulimia, history of seizures or unconsciousness; pregnancy or lactation; history of bipolar disorder; current major depressive disorder, use of any antidepressant medication; use of monoamine oxidase inhibitors within two weeks of study start; known allergies to the pharmacotherapies; suicidal or psychotic symptoms; recent treatment for drug or alcohol use; physical impairment so severe that the patient is unable to participate in a program of moderate physical activity.

#### *1.3. Trial Protocol to 12 Weeks*

At the baseline session, participants completed a physical exam, a epidemiological questionnaire, and a smoking-history assessment. Participants provided blood for a metabolic screen and CO to confirm smoking status. Participants attended group counseling sessions (see details below), and received combined sustained release bupropion and nicotine replacement therapy, which included regular meetings with medical staff (see details below).

At the 12 week assessment, CO-verified 7 day point prevalence abstinence (abstinence) was assessed. Both cessation trials included chronic treatment beyond 12 weeks; the current analysis used baseline and selected trial data to 12 weeks.

#### *1.4. Behavioral Intervention*

Group therapy consisted of five group sessions (once in Week 1, twice in week 3 (with quit day the date of the second session in week 3), once in week 5, and once in either week 11 (NCT00087880) or week 8 (NCT00086385), with individualized quit planning and cessation effort review. In preparation for chronic treatment, NCT00086385 participants randomized into two of four chronic treatment arms received additional counseling sessions in weeks 10 and 12.

#### *1.5. Medication*

Medication was combined sustained release bupropion (BUP) and nicotine replacement therapy (NRT). Participants received 12 weeks of BUP starting in week 1 (initially 150 mg/day, increasing to 150 mg b.i.d. on day four for NCT00087880 participants and at the beginning of week 2 for NCT00086385 participants). NRT was provided to participants to begin in week 2 on the quit day. NCT00087880 participants were provided with 6 weeks of 21 mg patch, 2 weeks of 14 mg patch and 2 weeks of 7 mg patch. NCT00086385 participants were provided with 10 weeks of 2 mg gum if smoking $< 25$ CPD, or 6 weeks of 4 mg gum followed by 4 weeks of 2 mg gum if smoking $\geq$ 25 CPD. Those using 2 mg gum who chewed $>$ 12 pieces a day or had difficulty maintaining abstinence were switched to 4 mg gum. In preparation for chronic treatment, NCT00086385 participants in two of four chronic treatment arms were instructed to taper their use of NRT gum beginning at week 8 and to discontinue NRT gum use by week 12. Participants met with medical staff in the same weeks they met for group counseling to receive instructions, ask questions, and relay concerns.

#### *1.6. Clinical Treatment Variables*

Clinical treatment variables indicating source RCT (Hall et al. 2011, 2009), instructions to taper from NRT from weeks 8 to 12 (Hall et al. 2009), and assignment of additional counseling sessions in weeks 10 and 12 (Hall et al. 2009) were created (see **Table 1**). These variables address differences in treatment across trials and among participants in the last month before the 12 week assessment.

### 2. Pharmacogenetic Study

We studied participants of two randomized clinical trials (Hall et al. 2011, 2009) who agreed to be in a Pharmacogenetic Study, and who provided a blood sample for DNA extraction, genotyping and genetic analysis of nicotine dependence, nicotine metabolism and clearance, and response to treatment (Bergen et al. 2013, 2014, 2015). Thus, Pharmacogenetic Study participants were interested in cessation treatment and contributing to related genetic research.

Analysis of Pharmacogenetic Study recruitment proportions from the two clinical trials and comparison of genetic study and clinical trial participants used Pharmacogenetic Study data and published RCT data (Hall et al. 2008, 2009, 2011; Hendricks et al. 2008). Similar proportions of RCT participants were recruited into (and withdrew from) the Pharmacogenetic Study from the two RCTs, suggesting recruitment of participants into the Genetic Study was not biased by RCT (**Supplementary Table 1**). Selected covariates and smoking measures were compared in the combined RCTs participant and the Pharmacoenetic Study participants (**Supplementary Table 2**); no significant differences were observed, suggesting that recruitment of participants into the Pharmacogenetic Study was not biased by these characteristics.

#### *2.1. Variables*

See above and the METHODS and **Table 1** in the main text for clinical, demographic, social, smoking, and genomic variables used in this analysis.

### 3. Analyses

Prediction of the urinary nicotine metabolite ratio and ancestry proportions in the Pharmacogenetic Study participants utilized novel genomic data generated for this analysis from existing DNA specimens.

#### *3.1. Genomic Data Analysis*

200 ng of genomic DNA were plated using Axiom 2.0 Reagent Kits and processed on the GeneTitan MC instrument. Analysis of the raw data was performed using Affymetrix Power tools (APT) v-1.16. Additional steps were

performed using SNPolisher to identify and select probe sets and high quality variants for downstream analysis. Quality control steps for samples included comparisons of self-reported and genomic gender and ancestry, detection of excessive heterozygosity (>0.20), genotype concordance among known duplicates, and removal of unexpected duplicates and related samples. Quality control steps for genetic variants included missingness >5% and deviation from Hardy Weinberg equilibrium ($p$<1E-10).

From N=454 DNA samples received, N=13 failed genotyping (11 samples due to a genotype rate threshold failure (<97%), one scan failure and one genotype clustering failure), and N=5 duplicate samples were removed, leaving N=438 participants with genome-wide data.

We used genome-wide imputation to harmonize genotypes across the studies prior to analysis. Alleles were reported on the forward strand and `conform-gt` was used to ensure consistency with the 1000 Genomes Phase 2 version 5a data files prepared for use with the Beagle imputation software. Beagle 5.2 was used to phase genotypes and impute ungenotyped or missing genotypes **?**. The resulting genotype dosages for variants typed on the Smokescreen Genotyping Array were imported into a Postgres database.

#### *3.1.1. Datasets used in Nicotine Metabolism Modeling*

As in Bergen et al. 2021, we used data from the Multiethnic Cohort to select and to train markers in statistical or machine learning models. In addition to performing the same approach of marker selection, model training and estimation within the Multiethnic Cohort, we enlarged the selection of variants to a total of four multiethnic datasets with measured NMR, and used the three additional multiethnic datasets for validation. These included: the Center for the Evaluation of Nicotine in Cigarettes (CENIC) study, which conducted studies on the effects of reduced nicotine cigarettes on smoking behaviors and exposures (Donny et al. 2015); the Hawaii Smokers Study, which studied urinary metabolites and the risk of lung cancer Derby et al. 2008, and Laboratory Studies of Nicotine Metabolism from three studies of labeled nicotine and cotinine (Swan et al. 2003; Dempsey et al. 2004; Swan et al. 2004), where we previously studied the genomics of plasma or saliva NMR Baurley et al. 2016) and developed a machine learning model of the NMR (Baurley et al. 2018) using variants at *CYP2A6* and at genes whose protein products regulate CYP2A6 activity.

#### *3.1.2. Variant Selection, Training, Validation and Prediction*

In each dataset we performed marker selection using a marginal scan including covariates (age, gender, BMI, and ancestry). Note that we used genomic ancestry as measured above instead of self-identified OMB race and ethnicity. We included the first 50 principal components to account for genetic relatedness among study participants. From each of the marginal scans we selected the top-ranked 200 variants and included the total (263 variants) in model training, together with age, gender, BMI, genomic ancestry.

To develop a predictive model of NMR, we took an ensemble based approach that leveraged a suite of machine learning algorithms. This suite consisted of partial least squares (Vinzi et al. 2010), projection pursuit (Huber 1985), elastic net (Zou and Hastie 2005), support vector machine (with a linear and radial basis function kernel) (Awad and Khanna 2015), gradient boosting machine (Natekin and Knoll 2013), and random forests (Breiman 2001). Each of these machine learning models was fit to the MEC data, treating the $Y_i$ (NMR measured on the $i$th subject) as the response variable. To explain the heterogeneity in the NMR in this analysis, we used a feature set consisting of age, gender, BMI, genetic ancestry, and the 263 prioritized markers arising from the marker nomination step. The R package `caret` was used to fit and train all of the models.

Once the process of training the models was complete, the ensemble model was constructed. In brief, the ensemble provides predictions by averaging the predictions of the individual sub-models. To examine the out of sample performance of our trained ensemble, we used it to predict the NMR for the subjects in the CENIC, HSS, and METS studies. Thus, the novel model predicting the uNMR has been externally validated with both uNMR and NMR measures (Baurley et al. 2021). Pharmacogenetic clinical and genotype data and the weights from the seven models were used to estimate the uNMR in Pharmacogenetic Study participants.

*3.2. Descriptive, Concurrent and Predictive Analyses*

See details of univariate association with abstinence in **Supplementary Table 3**. See smoking measures stratified by covariates in **Supplementary Table 4**. See covariate effects in two covariate models evaluating association of the uNMR with smoking measures in **Supplementary Tables 5** and **6**, and association of smoking measures and the uNMR with abstinence in **Supplementary Tables 7** and **8**.

*3.3. Strengths and Limitations*

Mean age and gender sex distributions of the Pharmacogenetic Study matched distributions seen in a contemporaneous hospital admissions dataset, and sex distributions of the Pharmacogenetic Study matched those of current or recent smokers in that dataset, measured using a conservative serum cotinine threshold (Benowitz et al. 2009). While African American, Asian and Hispanic participants in the Pharmacogenetic Study were underrepresented relative to proportions in the hospital population of smokers (Benowitz et al. 2009), minority representation in the Pharmacogenetic Study was four times the mean minority representation in contemporaneous NIH-sponsored trials (Ma et al. 2021).

The level of nicotine dependence in the Pharmacogenetic Study is higher than in contemporaneous (2000s) or later (2010s) populations because of cessation trial smoking behavior-related ascertainment criteria, and increases in light and moderate smoking and decreases in heavy smoking ($\geq$20 CPD) (Sakuma et al. 2016). Historical reductions in smoking intensity suggest cessation research may benefit from accurate measures of nicotine consumption.

Pharmacogenetic Study participants were recruited from a sample of treatment-seeking smokers already screened for medical and psychiatric contraindications. The three most common exclusions among N=2,799 individuals excluded were current use of antidepressants (23%), smoking <10 CPD (14%), and medical contraindications (11%) (Hall et al. 2008). However, potential RCT participants were not excluded due to some common histories; a history of DSM-IV Major Depressive Disorder was common (28%) in the RCTs, while only 0.5% of exclusions were due to a current diagnosis of Major Depressive Disorder (Hall et al. 2008).

In this analysis we approached variable analysis in a conservative manner (pre-planned analyses and dichotomous stratification of most covariates), and future research might use alternative approaches. We lacked socioeconomic variables other than education. The most profound limitation was lack of information on use of mentholated cigarettes, subject to targeted marketing to African Americans, and associated with reduced quit ratios in African Americans (Smith et al. 2020).

#### *3.4. Data Availability*

The datasets generated during and/or analysed during the current study are available from the corresponding author on reasonable request.

*3.5. Acknowledgments*

We thank the participants, Principal Investigators, co-Investigators and staff of the RCTs and the Pharmacogenetic Study. The National Institute on Drug Abuse funded the RCTs (R01 DA02538, K05 DA016752, K23 DA018691, and P50 DA09253), and the Pharmacogenetic Study (U01 DA020830). We thank Denise Nishita, Lisa Jack and Gary Swan (site-PI), Pharmacogenetics of Nicotine Addiction Treatment Consortium at SRI International (U01 DA020830, MPI: NL Benowitz, RF Tyndale and C Lerman), for their essential contributions that enabled this study.

The METS were supported by: the National Institute on Drug Abuse Pharmacokinetics and Pharmacodynamics of Nicotine (DA002277, PI: Neal L Benowitz), Young Adult Substance Use—Predictors and Consequences (DA003706, PI: Hy Hops), Pharmacokinetics of Nicotine in Twins (DA011170, PI: Gary E Swan), Pharmacogenetics of Nicotine Addiction Treatment Consortium (DA020830, PI: Neal L Benowitz; MPI: Rachel F Tyndale, and Caryn Lerman); and, by the Tobacco-Related Disease Research Program of the University of California: Nicotine Metabolism in Families (7PT-2004, PI: Neal L Benowitz).

We acknowledge BioRealm LLC team (`https://biorealm.ai`) for supporting project workflows and computation, and, IBX (`http://ibx.bio`) for DNA sample processing.

### 4. Tables

**Table 1:** RCT and Genetic Study Recruitment

| Study | NCT00087880* | | NCT00086385** | |
|---|---|---|---|---|
| Stage | Treatment | Genetic | Treatment | Genetic |
| Telephone screen | 2,508 | n/a | 2,314 | n/a |
| Excluded | 1,221 (48.7%) | n/a | 1,210 (52.3%) | n/a |
| Caller terminated | 119 (4.8%) | n/a | 119 (5.1%) | n/a |
| Invited to orientation | 1,168 | n/a | 985 | n/a |
| Attended | 607 (52.0%) | n/a | 620 (62.9%) | n/a |
| Eligible agreed attend baseline | 480 (79.1%) | n/a | 518 (83.6%) | n/a |
| Medical exclusion | 31 (6.5%) | n/a | 60 (11.6%) | n/a |
| Psychiatric exclusion | 21 (4.4%) | n/a | 29 (5.6%) | n/a |
| Eligible to participate | 428 (89.2%) | n/a | 429 (82.8%) | n/a |
| Decided not to participate | 21 (4.9%) | n/a | 26 (6.1%) | n/a |
| Deceased prior randomization | 1 | n/a | 1 | n/a |
| Enrolled in RCT | 406 | n/a | 402 | n/a |
| Invited | n/a | 332 (81.8%)$^{\alpha}$ | n/a | 344 (85.6%)$^{\alpha}$ |
| Consented | n/a | 226 (68.1%)$^{\beta}$ | n/a | 240 (69.8%)$^{\beta}$ |
| Remained | n/a | 220 (97.3%)$^{\gamma}$ | n/a | 236 (98.3%)$^{\gamma}$ |

*Hall et al. 2011. **Hall et al. 2009. $^{\alpha}$Invited into Genetic Study/Enrolled in RCT ($\chi^2$=2.13, $P$=.14). $^{\beta}$Consented/Invited ($\chi^2$=0.23, $P$=.63). $^{\gamma}$Remained after withdrew from Genetic Study (Fisher $P$=0.53).

**Table 2:** Genetic Study and RCT Sociodemographics and Smoking

| Characteristic | Genetic Study M (SD) or N (%) | Combined RCTs M (SD) or N (%) | Test, $P$ |
|---|---|---|---|
| N | 456 | $810^{\alpha,\beta}$ | |
| Age years | 49.5 (11.1) | 48.7 $(11.3)^{\beta}$ | $t$=1.27, $P$=.90 |
| Education | | | $\chi2$=0.42, $P$=.52 |
| $<$ College grad. | 224 (49.3%) | 409 $(51.3\%)^{\alpha}$ | |
| $\geq$ College grad. | 230 (50.7%) | 389 (48.7%) | |
| Female | 189 (41.4%) | 321 $(39.6\%)^{\alpha,\beta}$ | $\chi2$=0.40, $P$=.53 |
| Race | 447 | est. from $\%^{\beta}$ | $\chi2$=0.35, $P$=.95 |
| African American | 33 (7.4%) | 65 (8%) | |
| Multiple | 42 (9.4%) | 73 (9%) | |
| Other | 29 (6.5%) | 48 (6%) | |
| White | 343 (76.7%) | 624 (77%) | |
| Hispanic ethnicity | | est. from $\%^{\beta}$ | |
| (Yes/No) | 30/415 (6.7%) | 49/761 (6%) | $\chi2$ = 0.23, $P$=.63 |
| CO | 20.9 (10.8) | 21.5 $(10.7)^{\beta}$ | $t$=-0.95, $P$=.17 |
| CPD | 19.4 (7.7) | 19.8 $(8.1)^{\beta}$ | $t$=-0.87, $P$=.19 |
| FTCD | 4.8 (2.1) | 4.9 $(2.0)^{\alpha,\beta}$ | $t$=-0.82, $P$=.21 |

$^{\alpha}$Hall et al. 2008. $^{\beta}$Hendricks et al. 2008.

**Table 3:** Participant Characteristics and Abstinence, $N = 456$

| Variable | Not Abstinent | Abstinent | $\chi^2$ or $t$ | 2-sided $P$ |
|---|---|---|---|---|
| NCT00086385 | 98 (51.3%) | 138 (52.1%) | 0.026 | .872 |
| NCT00087880 | 93 (48.7%) | 127 (47.9%) | | |
| TAPER Yes | 50 (26.2%) | 62 (23.4%) | 0.464 | .496 |
| TAPER No | 141 (73.8%) | 203 (76.6%) | | |
| SESSION Yes | 39 (20.4%) | 57 (21.5%) | 0.079 | .778 |
| SESSION No | 152 (79.6%) | 208 (78.5%) | | |
| AGE $<$50 | 79 (41.4%) | 108 (40.8%) | 0.017 | .896 |
| AGE $\geq$50 | 112 (58.6%) | 157 (59.3%) | | |
| Female | 87 (45.5%) | 102 (38.5%) | 2.279 | .131 |
| Male | 104 (54.5%) | 163 (61.5%) | | |
| $<$Col. grad. | 102 (53.4%) | 122 (46.0%) | 2.776 | .096 |
| $\geq$Col. grad. | 87.0 (45.5%) | 143 (54.0%) | | |
| African Amer. | 21 (11.0%) | 12 (4.5%) | 7.5 | .006 |
| Multiple | 19 (9.9%) | 23 (8.7%) | 0.595 | .441 |
| Modified Other | 12 (6.3%) | 17 (6.4%) | 0.090 | .807 |
| White | 134 (70.2%) | 209 (78.9%) | reference | |
| Hispanic | 12.0 (6.3%) | 18.0 (6.8%) | 0.033 | .856 |
| Non-Hispanic | 173 (90.6%) | 242 (91.3%) | | |
| BMI $<$25 kg/m$^2$ | 91 (47.6%) | 110 (41.5%) | 1.293 | .256 |
| BMI $\geq$25 kg/m$^2$ | 97 (50.8%) | 146 (55.1%) | | |
| African ancestry | 0.13 (0.28) | 0.05 (0.18) | -3.14 | .002 |
| Asian ancestry | 0.07 (0.19) | 0.09 (0.25) | 0.74 | .456 |
| CO | 21.7 (10.5) | 20.4 (11.1) | -1.27 | .203 |
| CPD | 20.5 (8.0) | 18.5 (7.4) | -2.71 | .007 |
| FTCD | 5.0 (2.0) | 4.6 (2.1) | -2.34 | .019 |
| TTFC | 2.0 (0.9) | 1.7 (1.0) | -2.83 | .005 |
| F_CPD | 1.5 (0.7) | 1.4 (0.7) | -1.24 | .215 |
| uNMR | 1.27 (0.41) | 1.19 (0.44) | -1.95 | .052 |

Missingness: Education, n=2; Race, n=9; Ethnicity, n=11; BMI, n=12; Genomic (uNMR and ancestry proportions) n=20; FTND, n=7; TTFC n=11; FTND CPD item, n=8.

**Table 4:** Covariates, Smoking Measures and uNMR

| | Age | | Gender | | Race | | | | Ethnicity | | BMI | | Education | |
|---|---|---|---|---|---|---|---|---|---|---|---|---|---|---|
| | <50 | ≥50 | F | M | Afr | Mult | Oth | Wh | H | NH | <25 | ≥25 | <Col. grad. | ≥Col. grad. |
| CO | 20.2 | 21.4 | 20.2 | 21.5 | 21.1 | 18.9 | 19.5 | 21.3 | 18.9 | 21.1 | 20.7 | 20.7 | 21.6 | 20.4 |
| CPD | 18.0 | 20.4 | 17.6 | 20.6 | 16.7 | 16.8 | 18.0 | 20.1 | 19.1 | 19.2 | 18.6 | 19.8 | 20.1 | 18.7 |
| FTCD | 4.73 | 4.80 | 4.53 | 4.93 | 4.53 | 4.50 | 4.44 | 4.88 | 4.27 | 4.79 | 4.61 | 4.87 | 4.98 | 4.55 |
| TTFC | 1.75 | 1.89 | 1.86 | 1.79 | 1.76 | 1.86 | 1.70 | 1.87 | 1.74 | 1.86 | 1.79 | 1.86 | 1.92 | 1.76 |
| FCPD | 1.32 | 1.53 | 1.34 | 1.52 | 1.23 | 1.14 | 1.27 | 1.51 | 1.34 | 1.44 | 1.37 | 1.49 | 1.45 | 1.43 |
| uNMR | 1.19 | 1.25 | 1.26 | 1.20 | 1.10 | 0.84 | 0.99 | 1.30 | 1.24 | 1.22 | 1.28 | 1.17 | 1.20 | 1.25 |

Covariates. Age (<50 and ≥50 years), Gender (F, Female; M, Male), Race (Afr=African American; Mult=Multiple; Oth=(Asian, Pacific Islander, Native American, and Other); Wh=White), Ethnicity (H=Hispanic, NH=Non-Hispanic), BMI=Body Mass Index (<25 and ≥25 kg/m$^2$), and Education (<College graduate, ≥College graduate). Smoking measures: CO (ppm), CPD=Cigarettes per day (continuous), FTND=FTCD Total, TTFC=FTCD Time-To-First-Cigarette item, FCPD=FTCD CPD item, and uNMR=Predicted natural log uNMR.

**Table 5:** Covariate Model 1, uNMR and Smoking Measures

| Variable | CO | CPD | FTND | TTFC | FCPD |
|---|---|---|---|---|---|
| | $\beta$ (SE) *P* | $\beta$ (SE) *P* | $\beta$ (SE) *P* | $\beta$ (SE) *P* | $\beta$ (SE) *P* |
| uNMR | 3.80 (1.29) .003 | 1.39 (0.89) .120 | 0.18 (0.25) .469 | 0.10 (0.12) .400 | 0.07 (0.08) .390 |
| Age | 0.90 (1.03) .387 | 2.35 (0.71) .001 | 0.09 (0.20) .469 | 0.15 (0.09) .100 | 0.20 (0.06) .001 |
| Gender | 1.57 (1.05) .135 | 2.74 (0.72) <.001 | 0.35 (0.20) .651 | 0.10 (0.09) .316 | 0.16 (0.06) .013 |
| Education | -1.45 (1.03) .160 | -1.59 (0.71) .026 | -0.46 (0.20) .079 | -0.17 (0.09) .063 | -0.04 (0.06) .527 |
| African American | 0.50 (2.04) .806 | -3.18 (1.41) .024 | -0.48 (0.39) .219 | -0.13 (0.18) .469 | -0.30 (0.12) .015 |
| Multiple | -0.22 (1.84) .907 | -1.39 (1.27) .273 | -0.27 (0.35) .443 | -0.12 (0.17) .482 | -0.19 (0.11) .089 |
| Other | -0.45 (2.19) .837 | -2.44 (1.51) .106 | -0.33 (0.42) .432 | 0.02 (0.20) .937 | -0.31 (0.13) .019 |
| Ethnicity | -2.41 (2.11) .254 | -0.44 (1.45) .765 | -0.54 (0.40) .182 | -0.14 (0.19) .454 | -0.05 (0.13) .649 |
| BMI | 0.14 (1.06) .894 | 1.13 (0.73) .123 | 0.26 (0.20) .206 | 0.08 (0.10) .418 | 0.13 (0.06) .041 |

Smoking measures are outcomes and the uNMR is the predictor. Covariate Model 1 adjusted for (reference): Age (<50 years), Gender (Female), Education (<Col. Grad.), OMB Race (White), OMB Ethnicity (Non-Hispanic) and BMI (<25 $kg/m^2$). Smoking Measures: CO (ppm), CPD=Cigarettes per day, FTCD=FTCD Total, TTFC=FTCD Time-To-First-Cigarette item, FCPD=FTCD CPD item and uNMR=predicted natural log uNMR.

**Table 6:** Covariate Model 2, uNMR and Smoking Measures

| Variable | CO | CPD | FTND | TTFC | FCPD |
|---|---|---|---|---|---|
| | $\beta$ (SE) *P* | $\beta$ (SE) *P* | $\beta$ (SE) *P* | $\beta$ (SE) *P* | $\beta$ (SE) *P* |
| uNMR | 3.88 (1.32) .004 | 0.82 (0.90) 0.364 | 0.06 (0.25) .830 | 0.07 (0.12) .585 | 0.02 (0.08) .815 |
| Age | 0.98 (1.03) .342 | 2.37 (0.70) <.001 | 0.06 (0.25) .829 | 0.15 (0.09) .113 | 0.21 (0.06) <.001 |
| Gender | 1.42 (1.05) .176 | 2.81 (0.72) <.001 | 0.38 (0.20) .057 | 0.12 (0.09) .200 | 0.16 (0.06) .009 |
| Education | -1.44 (1.03) .166 | -1.67 (0.71) .019 | -0.44 (0.20) .027 | -0.16 (0.09) .096 | -0.05 (0.06) .461 |
| African ancestry | -0.87 (2.37) .713 | -4.34 (1.63) .008 | -0.48 (0.45) .289 | 0.06 (0.21) .764 | -0.42 (0.14) .004 |
| Asian ancestry | 0.50 (2.51) .842 | -4.81 (1.71) .005 | -0.96 (0.48) .045 | -0.28 (0.23) .218 | -0.52 (0.15) .001 |
| BMI | 0.22 (1.06) .838 | 1.06 (0.72) .143 | 0.22 (0.20) .285 | 0.05 (0.10) .584 | 0.13 (0.06) .047 |

Smoking measures are outcomes and the uNMR is the predictor. Covariate Model 2 adjusted for (reference): Age (<50 years), Gender (Female), Education (<Col. Grad.), African ancestry, Asian Ancestry and BMI (<25 kg/$m^2$). Smoking Measures: CO (ppm), CPD=Cigarettes per day, FTCD=FTCD Total, TTFC=FTCD Time-To-First-Cigarette item, FCPD=FTCD CPD item, and uNMR=predicted natural log uNMR.

**Table 7:** Covariate Model 1, Smoking Measures, uNMR and Abstinence

| Variable | CO | CPD | FTND | TTFC | FCPD | uNMR |
|---|---|---|---|---|---|---|
| | $\beta$ (SE) $P$ | $\beta$ (SE) $P$ | $\beta$ (SE) $P$ | $\beta$ (SE) $P$ | $\beta$ (SE) $P$ | $\beta$ (SE) $P$ |
| Smoking Measure | -0.01 (0.01) .193 | -0.05 (0.01) <.001 | -0.12 (0.05) .012 | -0.31 (0.10) .003 | -0.29 (0.15) .057 | -0.58 (0.25) .023 |
| Age | 0.05 (0.20) .823 | 0.14 (0.20) .502 | 0.03 (0.20) .860 | 0.07 (0.20) .713 | 0.08 (0.20) .681 | 0.05 (0.20) .823 |
| Gender | -0.18 (0.20) .381 | -0.33 (0.21) .105 | -0.25 (0.20) .219 | -0.24 (0.20) .243 | -0.25 (0.20) .212 | -0.18 (0.20) .381 |
| Education | 0.33 (0.20) .096 | 0.25 (0.20) .209 | 0.26 (0.20) .186 | 0.27 (0.20) .175 | 0.31 (0.20) .121 | 0.33 (0.20) .096 |
| African American | -1.09 (0.40) .007 | -1.16 (0.40) .004 | -1.05 (0.40) .008 | -1.05 (0.40) .009 | -1.07 (0.40) .007 | -1.09 (.40) .007 |
| Multiple | -0.45 (0.35) .206 | -0.34 (0.34) .322 | -0.29 (0.34) .393 | -0.30 (0.34) .378 | -0.31 (0.34) .364 | -0.45 (0.35) .210 |
| Other | -0.38 (0.42) .367 | -0.26 (0.41) .514 | -0.17 (0.40) .668 | -0.13 (0.40) .740 | -0.22 (0.41) .586 | -0.38 (0.42) .367 |
| Ethnicity | 0.18 (0.41) .665 | 0.07 (0.41) .850 | 0.02 (0.41) .947 | 0.05 (0.40) .906 | 0.07 (0.40) .851 | 0.18 (0.41) .665 |
| BMI | 0.27 (0.21) .184 | 0.38 (0.21) .063 | 0.36 (0.20) .077 | 0.36 (0.20) .082 | 0.36 (0.20) .074 | 0.27 (0.21) .184 |

Abstinence is outcome and smoking measures are predictors. Covariate Model 1 adjusted for (reference): Age (<50 years), Gender (Female), Education (<Col. Grad.), Race (White), Ethnicity (Non-Hispanic) and BMI (<25 kg/m$^2$). Smoking Measures: CO (ppm), CPD=Cigarettes per day, FTCD=FTCD Total, TTFC=FTCD Time-To-First-Cigarette item, FCPD=FTCD CPD item and uNMR=predicted natural log uNMR.

**Table 8:** Covariate Model 2, Smoking Measures, uNMR and Abstinence

| Variable | CO | CPD | FTND | TTFC | FCPD | uNMR |
|---|---|---|---|---|---|---|
| | $\beta$ (SE) *P* | $\beta$ (SE) *P* | $\beta$ (SE) *P* | $\beta$ (SE) *P* | $\beta$ (SE) *P* | $\beta$ (SE) *P* |
| Smoking Measure | -0.01 (0.01) .171 | -0.05 (0.01) <.001 | -0.12 (0.05) .014 | -0.29 (0.10) .005 | -0.28 (0.15) .063 | -0.54 (0.26) .039 |
| Age | 0.04 (0.20) .828 | 0.14 (0.20) .478 | 0.04 (0.20) .840 | 0.08 (0.20) .708 | 0.09 (0.20) .658 | 0.05 (0.20) .792 |
| Gender | -0.21 (0.20) .307 | -0.32 (0.21) .118 | -0.24 (0.20) .244 | -0.23 (0.20) .263 | -0.24 (0.20) .241 | -0.17 (0.20) .406 |
| Education | 0.27 (0.20) .171 | 0.22 (0.20) .118 | 0.24 (0.20) .232 | 0.25 (0.20) .212 | 0.28 (0.20) .164 | 0.30 (0.20) .138 |
| African ancestry | -1.36 (0.46) .003 | -1.57 (0.47) .001 | -1.41 (0.47) .003 | -1.35 (0.47) .004 | -1.45 (0.48) .002 | -1.49 (0.47) .002 |
| Asian ancestry | 0.25 (0.46) .590 | 0.02 (0.47) .966 | 0.15 (0.46) .740 | 0.17 (0.46) .704 | 0.12 (0.20) .795 | -0.10 (0.50) .834 |
| BMI | 0.35 (0.20) .083 | 0.41 (0.21) .048 | 0.38 (0.20) .062 | 0.37 (0.20) .070 | 0.38 (0.20) .058 | 0.31 (0.21) .136 |

Abstinence is outcome and smoking measures are predictors. Covariate Model 2 adjusted for (reference): Age (<50 years), Gender (Female), Education (<Col. Grad.), African ancestry, Asian Ancestry and BMI (<25 kg/m$^2$). Smoking Measures: CO (ppm), CPD=Cigarettes per day, FTCD=FTCD Total, TTFC=FTCD Time-To-First-Cigarette item, FCPD=FTCD CPD item, and uNMR=predicted natural log uNMR.
